# Distribution of SARS-CoV-2 RNA Signal in a Home with COVID-19 Positive Occupants

**DOI:** 10.1101/2020.11.30.20234393

**Authors:** Juan P. Maestre, David Jarma, Cesca Yu, Jeff Siegel, Sharon Horner, Kerry A. Kinney

## Abstract

Although many COVID-19 patients quarantine and recover at home, the dispersal of SARS-CoV-2 onto surfaces and dust within the home environment remains poorly understood. To investigate the distribution and persistence of SARS-CoV-2 in a quarantine home, samples were collected from a household with two confirmed COVID-19 cases (one adult and one child). Home surface swab and dust samples were collected two months after symptom onset (and one month after symptom resolution) in the household. The strength of the SARS-CoV-2 molecular signal in fomites varied as a function of sample location, surface material and cleaning practices. Notably, the SARS-CoV-2 RNA signal was detected at several locations throughout the household although cleaning appears to have attenuated the signal on many surfaces. Of the 24 surfaces sampled, 46% were SARS-CoV-2 positive at the time of sampling. The SARS-CoV-2 concentrations in dust recovered from floor and HVAC filter samples ranged from 10^4^-10^5^ N2 gene copies/g dust. While detection of viral RNA does not imply infectivity, this study confirms that the SARS-CoV-2 RNA signal can be detected at several locations within a COVID-19 quarantine home and can persist after symptoms have resolved. In addition, the concentration of SARS-CoV-2 (normalized per unit mass of dust) recovered in home HVAC filters may prove useful for estimating SARS-CoV-2 airborne levels in homes.

## Introduction

SARS-CoV-2 transmission inside buildings remains a significant concern as the COVID-19 pandemic continues to surge worldwide. While studies of SARS-CoV-2 contamination in buildings have focused on locations with outbreaks or medical facilities treating critical COVID-19 patients, most individuals with COVID-19 spend their recovery period quarantined at home. Reported transmission rates of SARS-CoV-2 in homes are relatively sparse and vary considerably (Li et al., 2020; Madewell et al., 2020; Qian et al., 2020; Wang et al., 2020b; Wu et al., 2020). However, household COVID-19 transmission may be more common than previously recognized with a secondary infection rate of 36% recently reported for a sample of 101 U.S. households with COVID-19 (Lewis et al., 2020). Unfortunately, the extent to which SARS-CoV-2 contaminates materials within a quarantine home remains poorly understood. Thus, it is difficult to establish if contaminated surfaces or dust reservoirs within a home affect the risk for transmission in households with COVID-19. This uncertainty is compounded by the complexity of airborne viral transport in the built environment as well as the effect of occupant cleaning practices on the dispersal and persistence of SARS-CoV-2 in homes. Here we report on the distribution of SARS-CoV-2 contamination detected on surface and dust samples collected from a quarantine household with two COVID-19 cases.

Both symptomatic and asymptomatic individuals with COVID-19 can emit SARS-CoV-2 into the air via breathing, coughing and talking (Ma et al., 2020; Pan et al., 2020; To et al., 2020). Viral shedding varies among individuals and can begin two days prior to symptom onset and persist for 14 days or even longer (Wölfel et al., 2020). Although young children with COVID-19 are often asymptomatic, SARS-CoV-2 viral loads in children can be high and subsequent infection of parents is possible (Lopez et al., 2020; Lu et al., 2020). One important pathway for COVID-19 transmission is the inhalation of aerosolized SARS-CoV-2 respiratory droplets generated by an infected individual (Allen and Marr, 2020; Prather et al., 2020). Larger respiratory droplets will often settle relatively quickly to the floor or other interior surfaces. However, local air currents as well as evaporation of droplets can lead to extended lifetimes. Smaller viral particles can remain airborne for hours or longer (Allen and Marr, 2020) with recent modeling results indicating that these droplets may also be significant drivers of person-to-person transmission (Augenbraun et al., 2020; Miller et al., 2020). Given that controlled laboratory studies indicate that SARS-CoV-2 aerosols may remain infectious for up to 3 hours or even longer in air (Van Doremalen et al., 2020), this is a concern for occupants sharing a home. Indeed, experimental studies have verified that a short aerosol release in one room of a house can distribute and eventually settle on surfaces throughout a house (Tang et al., 2020). While increasing ventilation is one of the major controls available to reduce airborne exposures to SARS-CoV-2 in buildings, this can be difficult to achieve in U.S. households where outdoor air ventilation rates are typically low (Bekö et al., 2016; Shrestha et al., 2019; Yamamoto et al., 2010). Finally, the temperature and relative humidity in homes can also affect the SARS-CoV-2 virus with longer viabilities and airborne survival times expected at lower temperatures and lower humidity levels (Biryukov et al., 2020; Guillier et al., 2020; Matson et al., 2020).

To our knowledge, SARS-CoV-2 contamination within a quarantine household has not been determined directly. However, measurements of SARS-CoV-2 distribution in other environments with COVID-19 occupants (e.g., healthcare facilities, quarantine units and cruise ships) provides insight as to the potential contamination that may be possible (Chia et al., 2020; Guo et al., 2020; Ong et al., 2020; Santarpia et al., 2020). In hospitals, for instance, SARS-CoV-2 has been detected on a variety of surfaces (floors, handrails, and soles of medical staff) as well as in short-term air samples collected near patients (Chia et al., 2020; Guo et al., 2020; Ye et al., 2020). The evidence suggests that some contaminated surfaces may serve as reservoirs for resuspension (Chia et al., 2020; Santarpia et al., 2020) and fecal aerosolization of SARS-CoV-2 during flushing of the toilet is another possibility (Elsamadony et al., 2021; Liu et al., 2020; Ma et al., 2020). In the more confined environment of a commercial cruise ship, Yamagishi et al. (2020) found that 10% of surface samples in case-cabins were SARS-CoV-2 positive but almost no positive detections were observed in common area surfaces or in air samples suggesting limited dispersal. Notably, numerous studies have detected SARS-CoV-2 on the surfaces of ventilation grates or air outlets in buildings with COVID-19 patients (Guo et al., 2020; Mouchtouri et al., 2020; Nissen et al., 2020; Santarpia et al., 2020). Recently, the SARS-CoV-2 virus was detected in 25% of the swab samples collected from a heating, ventilation and air conditioning (HVAC) system in one hospital (Horve et al., 2020), and in 36.8% of vent openings and 89% of HVAC filter dust samples in a COVID-19 ward (Nissen et al., 2020). The fact that the SARS-CoV-2 virus can be recovered from HVAC filter dust is not too surprising given that previous research has demonstrated that filters from central HVAC systems can serve as long-term spatially integrated samplers of the indoor environment. The filter forensics approach has been used for the assessment of particle-bond contaminants that accumulate in the dust collected on HVAC filters (Bi et al., 2018; Givehchi et al., 2019; Maestre et al., 2018; Noris et al., 2011) including viruses (Goyal et al., 2011; Prussin et al., 2016). When this approach is combined with HVAC parameters such as flowrate through the filter and usage time, it is possible to quantitatively estimate the time-averaged indoor concentrations of the particle-bound contaminants (Givehchi et al., 2019; Haaland and Siegel, 2017). This approach has not yet been used to estimate SARS-CoV-2 airborne concentrations but this could be possible if the SARS-CoV-2 concentrations in home HVAC filter dust were known.

In addition to investigating the dispersal of SARS-CoV-2 in home environments, the potential for SARS-CoV-2 to persist on materials and dust reservoirs within homes is an important consideration. Fundamental laboratory studies have demonstrated that SARS-CoV-2 can survive on paper for up to 3 hours, treated-wood and other low porosity surfaces for up to 24 hours (Aboubakr et al., 2020; Chin and Poon, 2020). Other studies have shown longer survivability times up to 28 days (Riddell et al., 2020) at 20°C for high porosity surfaces, such as paper and banknotes. Furthermore, some viruses such as Influenza A can remain infective upon resuspension from surfaces (Asadi et al., 2020).

Cleaning of contaminated surfaces with disinfectants is expected to mitigate the spread of SARS-CoV-2 in hospitals and other environments including homes (Hirotsu et al., 2020; Kampf et al., 2020; Ong et al., 2020). Beyond the frequency at which surfaces are sanitized, the effectiveness of cleaning agents vary widely (Sanekata et al., 2010). Because SARS-CoV-2 is an enveloped virus, surfactants, such as soapy water and other household cleaners, are expected to lyse the viral membrane and eliminate the infectivity of the virus on surfaces (Jahromi et al., 2020).However, the persistence of SARS-CoV-2 on surfaces is likely affected by the active ingredient in a given cleaner which, for EPA approved cleaners, includes quaternary ammonium salts, sodium hypochlorite, hydrogen peroxide among several others (Sanekata et al., 2010; Tuladhar et al., 2012b).

Investigating SARS-CoV-2 in homes and other built environments is crucial since the spread of SARS-CoV-2 is influenced not only by occupant behavior but also by the characteristics of the buildings themselves. While a substantial number of studies have investigated hospitals, quarantine units, restaurants, and cruise ships, additional research is needed in homes where many COVID-19 patients recover. The objective of this study was to determine the spatial distribution of the SARS-CoV-2 RNA signal in a quarantine home one month after two household members recovered from COVID-19. In addition to examining surface samples, we also quantify the viral signal in dust and surface swab samples from the home environment, providing the first concentration level (N2 gene copy numbers/g dust) for HVAC and floor dust samples for comparison with other built environments.

## Methods

Two members of a family in a home environment study (i.e., a parent (participant 1, P1) and a child (participant 2, P2)) experienced COVID-19 symptoms (CDC, 2020) that were confirmed with a positive COVID-19 test. The family remained quarantined in their home until symptoms resolved one month after symptoms began. The parent agreed to use a researcher-supplied home sampling kit to obtain samples of the home environment as well as to answer an IRB-approved survey on COVID symptoms and home environment management. All home environment samples were collected by the parent approximately one month after COVID-19 symptoms had resolved in the household.

### Sample Collection

A total of 22 swab samples were collected from a variety of surfaces across the home as well as from the surface of the HVAC filter. A single phosphate-buffered saline Tween-20 (PBST) wetted swab (Floq Swab, Copan, Murrieta CA) was used to swab each of the surfaces for 20 seconds. When possible, a 929 cm^2^ (1ft x1ft) area was swabbed. In other cases, where the sampling area was difficult to estimate (e.g., door handles), the entire available surface was swabbed. For the home HVAC filter, a 32.26 cm^2^ area was swabbed. Dust samples were also collected from the master bedroom floor and from the home HVAC filter via a handheld vacuum cleaner (Eureka, Medford, MA, USA). For each vacuum sample, a new vacuum thimble was inserted into a clean thermoset plastic nozzle (Indoor Biotechnologies, Charlottesville VA) attached to the hand-held vacuum cleaner. For floor samples, a 929 cm^2^ (1ft x1ft) area was vacuumed for one minute. For HVAC filter dust samples, the whole filter area (2064.5 cm^2^; 16 inch x 20 inch) was vacuumed for one minute. All samples were transported on ice and stored at −20 °C in the laboratory until extraction which occurred within 5 days of sample collection.

### Nucleic acids extraction and RT-qPCR

For total nucleic acids extraction, the MagMAX™ Total Nucleic Acids extraction kit (ThermoFisher Eugene, Oregon, USA) was used in combination with the KingFisher (ThermoFisher) nucleic acid extractor. At the beginning of sample processing, the dust cake from vacuum samples and the swabs were transferred to the extraction kit bead beating tubes (ThermoFisher) and processed per manufacturer’s instructions.

The concentration of SARS-CoV-2 in RNA extracts was determined in triplicate on the ViiA7 Real-Time PCR System (ThermoFisher). RT-qPCR utilized the CDC nCOV_N2 primer/probe and CDC nCOV_N1 primer/probe set (Lu et al., 2020) (Integrated DNA Technologies). Standard curves were developed using the 2019-nCoV_N_Positive Control (Integrated DNA Technologies, Coralville, USA). Samples were analyzed using the TaqMan™ Fast Virus 1-Step Master Mix in 20-µl reactions run at 50 °C for 5 min, 95 °C for 20s, followed by 40 cycles of 95 °C for 15 s and 60 °C for 60 s per the manufacturer’s recommendations. The limit of detection was established at 82 N1 gene copies recovered per sample, and 56 N2 gene copies recovered per sample (see Supplemental Information). To ensure the quality of the results, all qPCR analyses were performed in triplicate and positive controls (synthetic SARS-CoV-2, IDT CDC) as well as negative controls (PCR grade water) were used in each RT-qPCR plate. Additionally, negative controls were included in the study and processed concurrently with the samples to account for background material and reagent contamination. All negative controls indicated no presence of SARS-CoV-2 RNA in the materials or reagents used. The bovine respiratory syncytial virus (BRSV) was utilized to evaluate viral recoveries from each of the matrices (swabs and dust) collected in this study as is done in other environmental studies (Gonzalez et al., 2020).

### Home Data

Both participants lived full-time in the case study home before the onset of COVID-19 symptoms, during the disease period, and after the recovery period when the samples were collected. The two bedroom 93 m^2^ (1,000 ft^2^) home located in Texas was built between 2008 and 2012, and had a central HVAC system with a Minimum Efficiency Reporting Value (MERV) 4 (fiberglass filament, Flanders E-Z Flow II) filter installed.

### Survey Data

Information regarding COVID-19 symptoms, cleaning practices, and surface materials (Table 1) were gathered via remote survey utilizing the Research Electronic Data Capture (REDCap) platform (Harris et al., 2019; Harris et al., 2009). For surfaces such as carpet and hard flooring, the frequency of vacuum and mopping were also reported. The survey was administered entirely remotely via a survey link sent to the participant’s phone, made possible with the REDCap platform.

**Table 1.** Locations sampled across the home, surface material type and cleaning regime

| Location | Surface | Type of sample | Material | Cleaning product* | Cleaning Regime | Surface Area Sampled (cm<sup>2</sup>) | Detection |
| --- | --- | --- | --- | --- | --- | --- | --- |
| Entrance | Interior Door knob | Swab | Metal | Cleaner 2 | Once per day or more | Unknown | Negative |
|  | Interior door trim | Swab | Wood | Not Cleaned | None | Unknown | Negative |
|  | Exterior door trim | Swab | Wood | Not Cleaned | None | Unknown | Negative |
| Living Room | Floor | Swab | Vinyl | Cleaner 1 | Once per day or more | 929 | Negative |
|  | Tv top surface | Swab | Plastic | Not Cleaned | None | Unknown | Positive |
|  | Couch | Swab | Vinyl | Not Cleaned | None | Unknown | Positive |
| Kitchen | Counters | Swab | Laminate | Cleaner 3 | Once per day or more | 929 | Negative |
|  | Dinner Table | Swab | Laminate | Cleaner 3 | Once per day or more | 929 | Negative |
|  | Refrigerator handle | Swab | Plastic | Cleaner 3 | Once per day or more | Unknown | Negative |
|  | Sink Handles | Swab | Plastic | Cleaner 3 | Once per day or more | Unknown | Negative |
| Master bedroom | Door knob | Swab | Metal | Cleaner 2 | Once per day or more | Unknown | Positive |
|  | Floor | Swab | Carpet | Vacuum | Once per day or more | 929 | Positive |
|  | Floor | Vacuumed dust | Carpet | Vacuum | Once per day or more | 929 | Positive |
| Second Bedroom | Door knob | Swab | Metal | Cleaner 2 | Once per day or more | Unknown | Negative |
|  | Floor | Swab | Carpet | Vacuum | Once per day or more | 929 | Positive |
| Bathroom | Floor | Swab | Vinyl | Cleaner 1 | Once per day or more | 929 | Negative |
|  | Sink handles | Swab | Metal | Cleaner 2 | Once per day or more | Unknown | Negative |
|  | Toilet seat | Swab | Plastic | Cleaner 2 | Once per day or more | Unknown | Negative |
|  | Toilet handle | Swab | Metal | Cleaner 2 | Once per day or more | Unknown | Positive |
| Portable items | Phone screen 1 | Swab | Glass | Cleaner 2 | Once per day or more | Unknown | Negative |
|  | Toy | Swab | Plastic | Cleaner 2 | Less than once per day | Unknown | Positive |
|  | Highchair | Swab | Plastic | Water | Once per day or more | 929 | Positive |
| HVAC | Filter | Swab | Fiberglass | Not Cleaned | None | 32 | Positive |
|  | Filter | Vacuumed dust | Fiberglass | Not Cleaned | None | 2064 | Positive |

\* See Table S1 for active ingredient

### Quantitative Filter Forensics

The quantitative filter forensics approach by Haaland and Siegel (2017) was used in this work (Table S2) to estimate the temporally and spatially integrated airborne concentration (C) of SARS-CoV-2 over the viral collection time period. The following parameters were used for the calculation: ***m*** was the mass of dust (g) collected in the HVAC filter, ***f*** was the concentration of SARS-CoV-2 (N2 gene copies/g) in the dust collected on the filter, ***η*** was the integrated particulate matter filtration efficiency of the MERV-4 filter, ***Q*** was the volumetric air flowrate (m^3^/h) through the filter (median for the summer season for the same geographical location, (Givehchi et al., 2019) and ***t*** was the runtime of the HVAC system (h) over the duration of the SARS-CoV-2 collection time -approximately one month-, median for the summer season for the same geographical location, (Givehchi et al., 2019). A few considerations that ought to be taken into account are: (1) our estimate does not account for the attenuation of the signal over time or the losses due to deposition, (2) it considers the estimate that approximately 5.5% of the viral signal is recovered through RNA extraction from the dust matrix (as estimated in this work via the spike and recovery tests for the surrogate BRSV virus), (3) the mass recovered by the participants was estimated to be 68% of that recoverable by trained researcher (based on previous experiments comparing researcher-collected samples to participant-collected samples), and (4) that approximately 27% of the accumulated dust can be recovered from the filter (Mahdavi and Siegel, 2020). Owing to the use of averaged parameters from a similar population of homes and analysis approaches, our estimate should be considered a scaling approach rather than a precise calculation of SARS-CoV-2 airborne concentration.

## Results

### Ventilation and Temperature Settings

The home was naturally ventilated one hour per day, in the early morning by opening one door. The HVAC temperature setting was kept at 23.9°C (75°F) day and night with the air conditioning system providing cooling during the summer months in this hot and humid region of Texas (average ambient temperature of 35°C (95°F) over the summer months). In our previous study in Texas (Bi et al., 2018), the average relative humidity in homes during the summer was 56.6% ± 5.2% (n=93, measured over one month). Even though indoor relative humidity varies as a function of temperature, outdoor relative humidity, occupancy and other building factors, this provides us with a reasonable estimate for homes in this study. Regarding the position of the HVAC filter, the low efficiency filter (MERV-4) was positioned vertically in the unit 3 feet above floor height. The HVAC unit was located inside a closet with a louvered door that was normally kept closed.

### SARS-CoV-2 on Surfaces across the Home

A total of 24 surfaces distributed across six spaces in the home were swabbed on the same day approximately one month after the symptoms of both participants had resolved. A total of 46% of the samples were found to be positive (Table 1, Figure 1). In some cases, a specific area was sampled (HVAC filter, counters, floors, and highchair) and the results are reported in N2 gene copies/cm^2^ (Fig. 2A), whereas in other cases when it was not feasible to normalize by area -or by weight-(e.g., swabbing doorknobs, handles, among others), the results are reported as N2 gene copies recovered per swab (Fig 2B).

**Figure 1.**
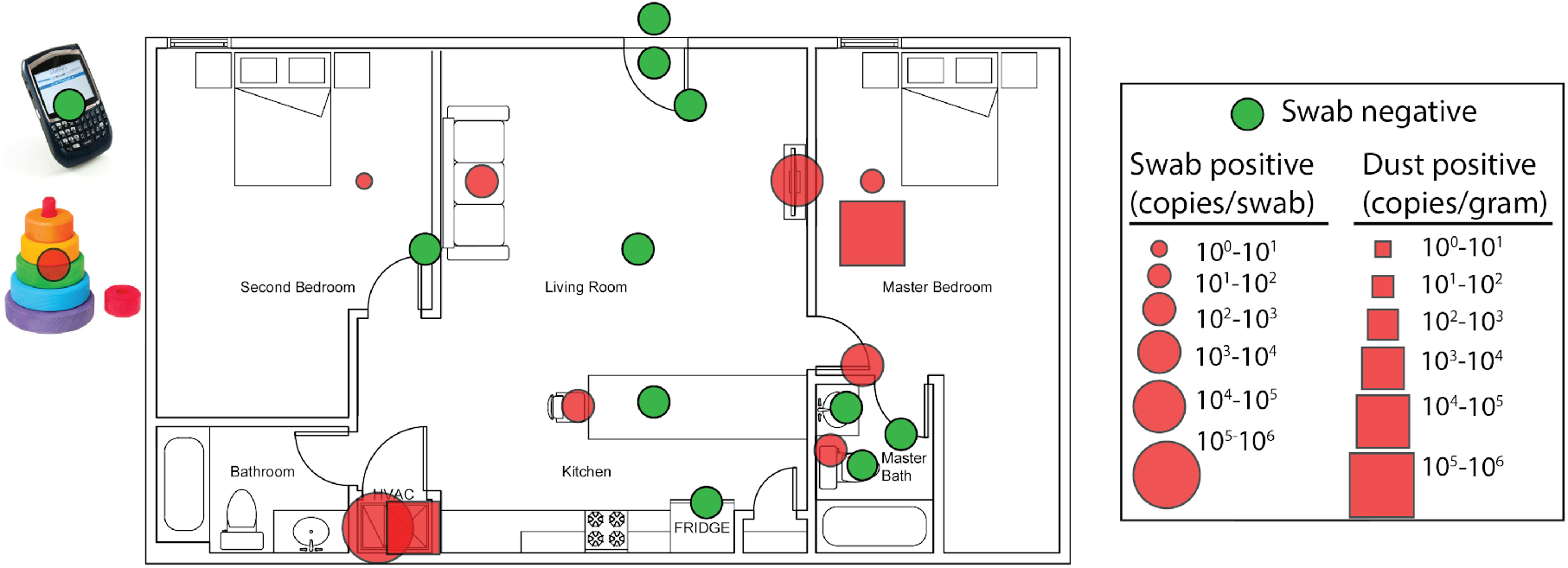
N2 gene copies recovered from samples. For visualization purposes only, results are illustrated on a generic two-bedroom floor plan that is typical of homes in the study area.

**Figure 2.**
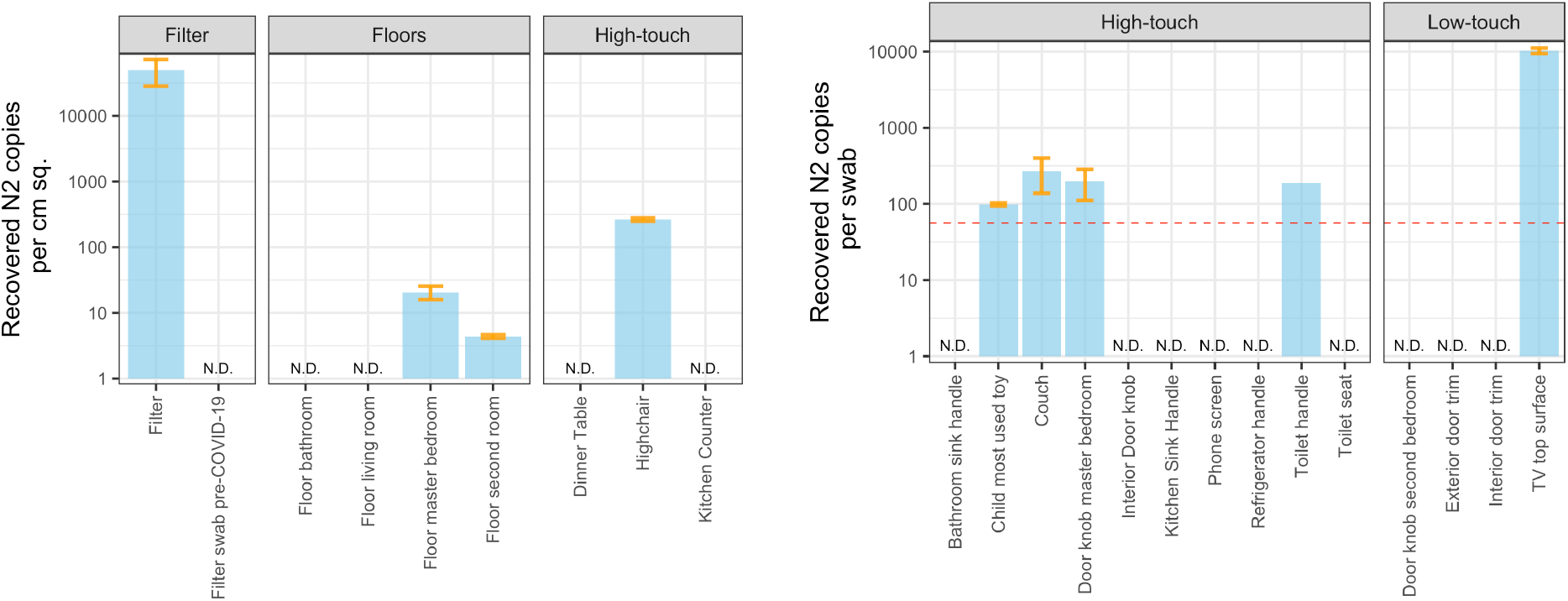
N2 gene copies recovered per swab across the fomites sampled. Left) Fomites sampled by area, allowing the results to be expressed in N2 gene copies/cm^2^. Right) Fomites sampled with no known areas, results presented in recovered copies per swab, red line represents the effective LOD. N.D.=Non-detects.

In the case of fomites where it was feasible to sample a specific area, the highest number of N2 gene copies/cm^2^ were recovered from the filter, whereas the floor in the master bedroom and the highchair yielded several order magnitude fewer copies/cm^2^. Samples gathered from the bathroom floor, living room floor and second bedroom floor, dinner table and kitchen counter yielded non-detects. In the carpet of the master bedroom, which was used as the primary bedroom by both occupants, a concentration of approximately 20 copies/cm^2^ was found. The highchair yielded concentrations one order of magnitude higher than the floor in the master bedroom. Interestingly, a concentration of 4.8 N2 gene copies/cm^2^ was measured in the second bedroom (not in use from the onset of symptoms through the sampling event). It is worth noting that vinyl flooring (bathroom and living room), cleaned with Cleaner 1 on a regular basis did not yield any signal, and neither did surfaces that were cleaned with Cleaner 3 such as the kitchen counter and the dinner table.

In the case of the results not normalized by area, the highest SARS-CoV-2 RNA signal was observed on the top of the television surface. The vinyl couch as well as the child’s toy, the toilet handle, and the doorknob to the master bedroom showed similar viral signal strengths (∼100 N2 gene copies recovered per swab). The surfaces in the bathroom did not yield any SARS-CoV-2 signal, except for the toilet handle. The remaining doorknobs (second bedroom and main door) and handles tested, including the kitchen sink handle and refrigerator handle, as well as the cell phone screen, did not yield any signal. Fig. S1 shows all swab samples as recovered copies per swab for direct comparison among all the samples without area normalization.

### Cleaning regime, products utilized and surface materials

Five methods for cleaning the home environment were used in the home (Table 1, Fig. 3). For vinyl flooring in the living room and kitchen, Cleaner 1 (active ingredient: glycolic acid) was used once per day, except in the case of the bathroom floor, where Cleaner 2 (active ingredient: alkyl dimethyl benzyl ammonium chloride) was generally utilized. For doorknobs, Cleaner 1 was utilized once per day. The rest of flooring in both bedrooms was carpeted. The floor in the main bedroom (the only one occupied at the time) was vacuumed once per day. In the kitchen, Cleaner 3 (active ingredient: sodium laureth sulfate) was employed to clean surfaces such as the counters, the dinner table, as well as the refrigerator and sink handles. In the bathroom, Cleaner 2 was used on all surfaces including the sink, toilet handle and toilet seat. Some portable objects were cleaned with Cleaner 2, including the participant’s phone, and the child’s toys, cleaned daily and 2-3 times per week respectively. The highchair was cleaned three times per day with water. Other surfaces, such as door trims, the couch, the TV, or the HVAC filter were not cleaned routinely.

**Figure 3.**
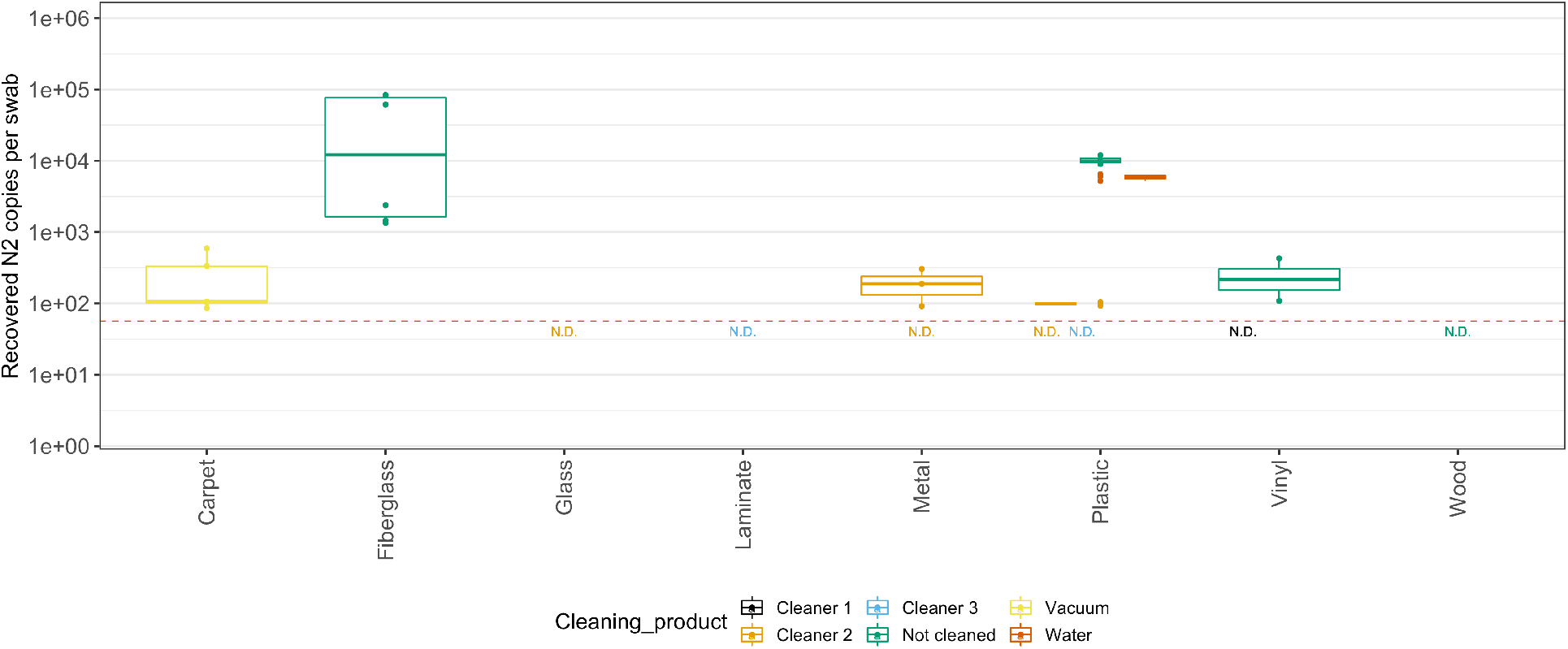
N2 gene copies recovered per swab from the fomites sampled as a function of surface material and cleaning product used on the surfaces. Red dashed line represents the effective LOD. Non-detects (N.D.) are represented by ½ of the LOD to facilitate interpretation of the figure.

The highest signal was recovered from the filter, made of fiberglass and never cleaned/disturbed by the participant until the sampling event, followed by the plastic materials (TV top and highchair), and carpet floor. Among the fomites that can be cleaned frequently, those that were vacuumed, not cleaned at all, or cleaned with water only, yielded higher SARS-CoV-2 signal than those cleaned with commercial cleaning products. Another factor that may affect the signal is the cleaning regime (Table 1, Fig. S2) with frequent cleaning of surfaces expected to deplete the signal.

### SARS-CoV-2 in vacuumed dust samples

The viral signal ranged from 10^5^ to 10^6^ N2 gene copies/g in the floor dust samples collected via the handheld vacuum (Fig. 4). The viral signal in the HVAC filter dust ranged from 10^4^ to 10^5^ copies/g of dust. The concentrations found in the dust from the carpeted master bedroom floor were significantly higher than those in the HVAC filter dust (Mann-Whitney, p-val<0.001). In both cases, the SARS-CoV-2 signal varied by at least an order of magnitude in the dust samples. As a negative control, vacuum HVAC filter dust samples collected a year prior to the COVID-19 pandemic were also tested and yielded no signal.

**Fig 4.**
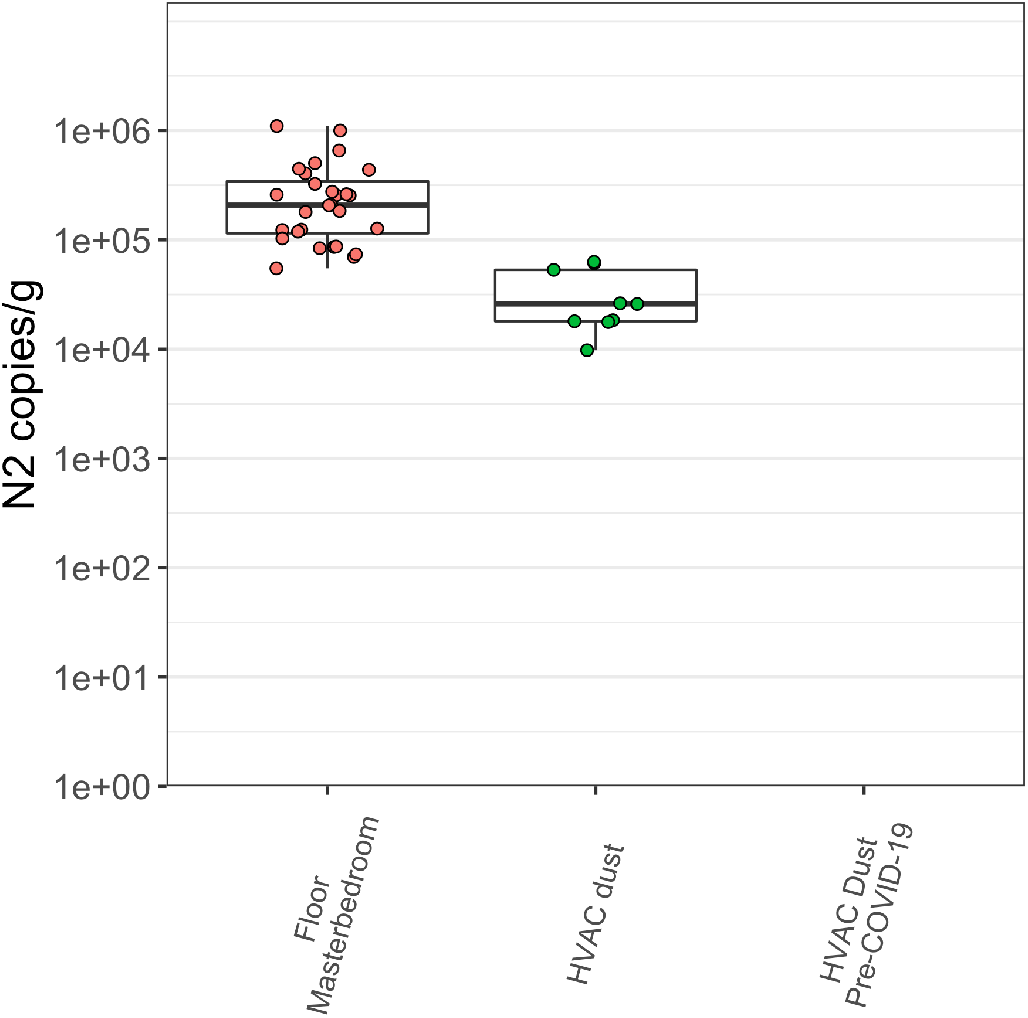
Concentration of N2 gene copies detected in vacuumed dust samples. Dust samples were aliquoted and replicates were measured to study the variability in the signal.

### N1-N2 Recovery comparison and estimation of viral recovery with BRSV virus

A subset of positive samples for the N2 assay were tested with the N1 assay to corroborate the results. In each case, all samples yielded positive signals, that were highly correlated (Pearson’s product-moment correlation, p-value = 0.002, rho =0.825), although N2 consistently yielded higher copies recovered than N1 (Figure S4).

It has been established that microbial RNA/DNA can be lost during the process of extraction (Iker et al., 2013) and by the matrix within which it is embedded (Zuo et al., 2013). To estimate the effect that matrix can have on viral recoveries (Gonzalez et al., 2020), the BRSV was used to estimate the viral recovery from swab and dust samples, In this work, when the RNA of a known amount of virus was extracted with no matrix-, approximately 30% of the signal was recovered (Fig. S3). When the same quantity of virus was spiked onto a swab (the same kind used in the current study), approximately 25% of the signal was recovered. However, in the case of BRSV spikes into dust samples, a median of approximately 6% was recovered from the filter dust and approximately 8% from the floor dust. As a reference, 7.6% recovery was observed for BRSV in wastewater samples and fungal recoveries from wipe samples have been reported to range from 10% to 25% Gonzalez et al., 2020; Yamamoto et al., 2011). Similarly, Yang et al. (2011) reported 50% recovery of the virus H1N1 spiked onto PTFE filters while Brown et al. (2020) could recover between 1% and 10% the foot-and-mouth disease virus in their liquid control with generally lower recoveries reported from surfaces, suggesting non-negligible losses. While it is known that different microorganisms, matrices, surface materials, and virus concentrations (Brown et al., 2020; Fabian et al., 2009; Zuo et al., 2013) can behave differently, the recovery values determined in the present work are within the range reported in other studies of viruses and other microorganisms. Still, the actual recovery of SARS-CoV-2 from many environmental matrices remains largely unknown.

## Discussion

Secondary SARS-CoV-2 infections can occur within households when a COVID-19 individual isolates at home (Grijalva et al., 2020; Lewis et al., 2020; Wang et al., 2020a; Wu et al., 2020). In fact, home related outbreaks can be common as evidenced in the study by Qian et al. (2020) where 254 of 318 outbreaks (79.9%) were found to have originated in the home indoor environment. Wang et al. (2020a) found that measures such as home ventilation and frequent cleaning of surfaces were protective against secondary infections. In this work, we detected SARS-CoV-2 (confirmed via N2 and N1 gene assays) across several locations within a home one month after the symptoms of the two COVID-19 positive occupants had subsided. The SARS-CoV-2 signal was found in at least one fomite in every room sampled, including in rooms not in active use. In addition, SARS-CoV-2 was detected in dust from floors and from the HVAC filter installed in the home. We found that 46% of the surfaces had detectable SARS-CoV-2 signal. In other studies (Chia et al., 2020), similar detection rates (∼40%) were found in rooms with patients in their first week of disease but the rates declined below 20% as the disease progressed. Many factors could have contributed to these observed differences. In our study, the strength of the viral signal seemed to vary with sample location, fomite surface material, occupant contact level, and cleaning practices.

Some of the SARS-CoV-2 positive surfaces, such as HVAC filters, floors, and the top of the TV, are common reservoirs for dust build-up and might be infrequently touched; however, others are high-touch surfaces such as doorknobs, tables and, handles. The viral signals recovered in the current study (median= 966 N2 gene copies recovered per swab sample) were lower than the median value of approximately 3500 N2 gene copies recovered per swab reported in biocontainment and quarantine units (Santarpia et al., 2020) although there was wide variability among sample types. Several relevant factors could explain these differences. First, it is possible that the viral signal in the study house attenuated as time passed after symptoms disappeared whereas the studies in hospitals and quarantine units were normally conducted in or near rooms with active COVID-19 cases. In the quarantine unit study by Santarpia et al. (2020), the rooms were negatively pressurized with >12 ACH whereas in homes, these ventilation levels are unlikely to be present in air conditioned homes that are closed most of the time. In the present study, the ACH during cooling was not measured although the home was naturally ventilated one hour per day. Finally, the home was cleaned frequently whereas in Santarpia et al. (2020), the authors mentioned frequent environmental cleaning but many details were not specified (cleaning regime, products), making the comparison difficult.

In some cases, the SARS-CoV-2 RNA signal can be normalized by sampling area, which may be a useful metric to compare to other studies. The highest concentrations of N2 gene copies/cm^2^ were recovered from the HVAC filter (average 43,000 N2 gene copies/cm^2^), which was in place throughout the period of illness and recovery. Other surfaces, such as the floor in the master bedroom and the second room, both carpeted and vacuum-cleaned, and the child’s highchair, cleaned with water, yielded signal but at lower concentrations (∼5, ∼20 and ∼125 copies/cm^2^, respectively). Comparing to samples gathered in hospitals, in Feng et al. (2021) 38 copies/cm^2^ were found at the patient’s bedside wall surface but lower concentrations in the toilet bowl and floor drainage (4 and 2 copies/cm^2^, respectively). In this study, samples gathered from the vinyl bathroom floor and living room floor, both cleaned with Cleaner 1, as well as the dinner table and kitchen counter (cleaned with Cleaner 3), yielded non-detects. It is noteworthy that 4.8 N2 gene copies/cm^2^ (nearly 100 N2 gene copies recovered per swab) were recovered in the unoccupied second bedroom. Many explanations are possible, including the possibility of tracking the virus in via walking, redistribution of viral particles via the HVAC system, or penetration of the virus from the occupied portion to the unoccupied bedroom via cracks and gaps in the doorframe. Evidence is mounting supporting the possibility of long travel distance of SARS-CoV-2 virus-laden particles in the built environment (Allen and Marr, 2020; Chen et al., 2020a; Morawska and Cao, 2020). Tang et al. (2020) found that aerosols released indoors dispersed and deposited across the open spaces, even within closets and cabinets with closed doors and drawers. Thus, the presence of viral signal in the not-in-use room carpeted floor is not unreasonable. Among the samples not normalized by area, the signal observed on the top of the television (nearly 10,000 N2 gene copies/swab) was significantly higher than the other samples. The static charge and lack of cleaning at this location leads to an accumulation of dust, suggesting that it could be a good reservoir to sample in home environmental studies, as reported elsewhere (Dunn et al., 2013).

Of particular note, the SARS-CoV-2 signal recovered by swabbing the HVAC filter in this study yielded an average viral signal of 38,815 N2 gene copies recovered per swab, higher than those recovered in the biocontainment and quarantine unit study. Horve et al. (2020) found SARS-CoV-2 viral copies in hospital HVAC pre-filters and filters at lower levels (∼ 450 cumulative copies across the pre-filters and two filters that were positive). Even though the levels found in the present work are higher, the direct comparison is difficult due to several aspects. First, home and hospital HVAC systems differ in their in their function; home systems provide greater air recirculation, whereas in hospital systems, outdoor air is mixed in to provide higher air exchange rates. The second main difference is the total volume of filtered air per surface sampled. Finally, the cumulative time the filters were in use may be different between the two systems. With the current available information, we are unable to determine the full influence of these significant factors. Nissen et al. (2020) found viral signal in air vents and filters 50 meters away from patients in a low relative humidity environment (approximately 30%), but they could not detect growth or infectivity, and hypothesized that the virus may have been inactive due to desiccation of the pathogen in the vents.

Recent studies have shown a variability in the survivability of the virus in surfaces, depending on the material, temperature, relative humidity, and light, with all of these factors playing an important role (Riddell et al., 2020; Van Doremalen et al., 2020; Wolff et al., 2005). Whereas one study showed that SARS-CoV-2 survives for up to 7 days in some materials (Aboubakr et al., 2020; Chin and Poon, 2020), other studies have shown longer survivability times up to 28 days (Riddell et al., 2020) in the dark, at 20 °C and 50% RH. Considering the time that had passed after the participants’ symptoms disappeared, the RH average estimate from our previous study in Texas (56.6% ± 5.2%, (Bi et al., 2018)), and the temperature setting for the study house (23.9 °C, 75 °F), we hypothesize that the viral signal detected may have been from inactive virus. However, additional studies that address infectivity of SARS-CoV-2 recovered from home surface and dust samples as a function of time, material properties and cleaning practices would be required to address this question.

Another factor that may affect the signal recovered from the built environment is the cleaning regime (Fig. S2) and the cleaning products used (Table 1, Table S1). In the present study, the frequent cleaning of floors and many of the high touch surfaces with cleaning products may explain the low to no signal found on many surfaces. Surfactants from household cleaners can break the viral membrane of the enveloped SARS-CoV-2 virus (Jahromi et al. 2020). For example, all surfaces in the bathroom cleaned regularly with Cleaner 2 (sink handle, toilet seat, and floor) did not yield any SARS-COV-2 signal, except in the case of the toilet handle suggesting that viral shedding in stool could have happened. Studies have found SARS-CoV-2 presence in stool samples up to 28 days after hospital admission (Xu et al., 2020), 6-10 days after negative nasopharyngeal swabs (Chen et al., 2020b). However, it is important to recognize that the viral signal detected in the toilet handle could also have settled there via transport from another area of the home. The persistence of the SARS-CoV-2 RNA signal on surfaces likely varies widely due to the active ingredient within the cleaner used. Quaternary ammonium, a compound found in Cleaner 2 in this study as well as in a majority of EPA recommended cleaners, has been shown to be effective against viruses (Shirai et al., 2000; Tuladhar et al., 2012a) such as the enveloped murine norovirus (MNV-1) and feline calicivirus (FCV) (Kennedy et al. 1995).

Ma et al. (2020) found that COVID-19 positive patients in early stages had breath emission rates estimated to range from 1.03×10^5^ to 2.25×10^7^ viruses per hour. It is likely that the concentrations would be lower towards the end of the recovery period as reported in sputum samples elsewhere (Wölfel et al., 2020). Taking into account the two COVID-19 positive dwellers, the length of their quarantine and ventilation mode (a door opened during one hour per day plus infiltration), it is reasonable that viral particles accumulated in the case study home. In this study, significant viral signals were recovered from the dust from both the main bedroom floor and the HVAC filter. Even though the floors of the study home were vacuumed frequently, the viral signal in the carpeted floor from the master bedroom was above 10^5^ N2 gene copies/ gram of dust, suggesting that the viral signal may be difficult to remove just by vacuuming. As indicated in Staudt et al. (2020), the highest viral loads can be found on floors and airborne aerosols can be formed from resuspension of settled dust or aerosols. In the present work, the signal found in the HVAC filter dust was an order of magnitude lower than that recovered from the floors. In both cases, the viral concentration in each dust sample type varied by at least an order of magnitude indicating that replicate analyses are needed to represent the heterogeneity of the dust. Solely for context, having not found other works providing SARS-COV-2 concentrations per gram of dust, the concentrations measured in this work are on the same order of magnitude or even higher than those found in stool from COVID-19 positive patients at the peak of their symptoms (Wölfel et al., 2020) but lower than those found in other cases (Lescure et al., 2020).

The virus is not expelled naked but attached to larger respiratory fluid particles, forming both droplets and aerosols that can be involved in transmission (Prather et al., 2020). Filters can remove airborne viruses, but their efficiency varies significantly. Low MERV filters are not very efficient at removing smaller particles with, for example, filters rated MERV 5 and below are reported to remove less than 25% of particles smaller than 10 µm (Azimi et al., 2014). As a result, only a fraction of viral aerosols may be retained in the filter in a single pass although our results suggest removal and accumulation can occur over time. Notably, we found a strong viral signal in the HVAC filter dust, located inside a closet with a closed louvered door. This indicates the viral-laden particles were at some point airborne (either after being expelled or after being resuspended). Low efficiency filters, frequently used because of their low cost, primarily act as a ‘roughing filter’, removing large particulate matter to protect the HVAC system. In order to enhance capture of viral-laden particles in a home (and potentially diminish the redistribution of viruses across the home), high MERV filters could be used.

This work indicates that it is possible to detect the previous presence of a viral shedding individual in the built environment via HVAC filter forensics and suggest this approach may be useful for SARS-CoV-2 monitoring in home environments. In addition, using the QFF methodology (Haaland and Siegel, 2017), we estimated an average integrated airborne SARS-CoV-2 concentration of 90 copies/m^3^ (see Table S2 for more details). This value can serve for future comparisons of concentrations of SARS-CoV-2 in the built environment and help building scientists and engineers as they strive to develop best practices in homes with COVID-19 positive occupants. Studies of the viral signal decay and infectivity in HVAC filter dust are warranted to further determine the usability and limits of this approach, and future work should include direct measurements of HVAC and extraction parameters to better characterize the integrated SARS-CoV-2 concentrations. Because HVAC filter dust represents a pooled sample, contributed to by all occupants in a building over an extended period of time, repeated monitoring could be used to identify spikes in measured viral concentrations in the dust. These spikes could indicate new or active infections and be used to guide additional isolation or ventilation practices to minimize the spread of the virus.

As the body of literature increases, it seems clear that in order to diminish viral loads and decrease the probablity of in-home secondary transmission in the built environment, efficient ventilation, high-efficiency filtration and frequent cleaning are important (Allen and Marr, 2020). While detection of viral RNA does not imply infectivity, this study confirms that the SARS-CoV-2 RNA signal can persist in a COVID-19 quarantine household for nearly a month following resolution of COVID-19 symptoms. In addition, several factors that may affect the distribution of SARS-CoV-2 across a home have been identified. The results indicate that cleaning can greatly reduce or eliminate the SARS-CoV-2 signal on surfaces. Also, even a low efficiency home filter is capable of capturing and retaining SARS-CoV-2. The detection of the SARS-CoV-2 RNA signal on infrequently touched surfaces indicates that airborne particles settle out of the air, potentially contaminating surfaces not in direct contact with COVID-19 positive individuals. As the COVID-19 pandemic continues, the SARS-CoV-2 transmission pathways continue to be widely debated. Homes are essential environments that warrant further study to better understand SARS-CoV-2 aerosols and fomites in the home environment where many COVID-19 individuals recover.

## Limitations of the Study

This study has several limitations. First, the distribution of the SARS-CoV-2 RNA signal across a single quarantine household was investigated at one time point two months after COVID-19 symptom onset. Additional quarantine homes during and after COVID-19 infections should be studied to determine the temporal course of SARS-CoV-2 distribution within homes as well as to investigate how different household factors affect this distribution across a wide variety of home types. Second, viral culturing was not performed to determine virus viability/infectivity; thus, this work only reports SARS-CoV-2 viral signal found independently of its viability. Future studies are needed to establish the whether there are infective viruses in samples obtained from home environments. Finally, the current study is based on the recovery of SARS-CoV-2 RNA from a range of sample types. It is well established that recovery of RNA from environmental samples is often attenuated by losses during the extraction process or retention within the sample matrix. Thus, future studies are warranted to investigate these effects as the recovery of SARS-CoV-2 from many complex sample matrices such as dust have not been established.

## Data Availability

Ct values and other supplemental data available upon request.

## Acknowledgements

This work was supported by Whole Communities—Whole Health, a research grand challenge at the University of Texas at Austin. In addition, the work that provided the basis for this publication was also supported by funding under an award TXHHU0046-18 with the U.S. Department of Housing and Urban Development. The substance and findings of the work are dedicated to the public. The authors and publisher are solely responsible for the accuracy of the statements and interpretations contained in this publication. Such interpretations do not necessarily reflect the views of the Government.

## References

Aboubakr HA, Sharafeldin TA, Goyal SM. Stability of SARS-CoV-2 and other coronaviruses in the environment and on common touch surfaces and the influence of climatic conditions: A review. Transboundary and Emerging Diseases 2020: 1–17.

Allen JG, Marr LC. Recognizing and controlling airborne transmission of SARS-CoV-2 in indoor environments. Indoor Air 2020; 30: 557–558.

Asadi S, Gaaloul ben Hnia N, Barre RS, Wexler AS, Ristenpart WD, Bouvier NM. Influenza A virus is transmissible via aerosolized fomites. Nature Communications 2020; 11: 4062.

Augenbraun BL, Lasner ZD, Mitra D, Prabhu S, Raval S, Sawaoka H, et al. Assessment and mitigation of aerosol airborne SARS-CoV-2 transmission in laboratory and office environments. Journal of Occupational and Environmental Hygiene 2020: 1–10.

Azimi P, Zhao D, Stephens B. Estimates of HVAC filtration efficiency for fine and ultrafine particles of outdoor origin. Atmospheric Environment 2014; 98: 337–346.

Bekö G, Gustavsen S, Frederiksen M, Bergsøe NC, Kolarik B, Gunnarsen L, et al. Diurnal and seasonal variation in air exchange rates and interzonal airflows measured by active and passive tracer gas in homes. Building and Environment 2016; 104: 178–187.

Bi C, Maestre JP, Li H, Zhang G, Givehchi R, Mahdavi A, et al. Phthalates and organophosphates in settled dust and HVAC filter dust of U.S. low-income homes: Association with season, building characteristics, and childhood asthma. Environment International 2018; 121: 916–930.

Biryukov J, Boydston JA, Dunning RA, Yeager JJ, Wood S, Reese AL, et al. Increasing temperature and relative humidity accelerates inactivation of SARS-CoV-2 on surfaces. MSphere 2020; 5.

Brown E, Nelson N, Gubbins S, Colenutt C. Environmental and air sampling are efficient methods for the detection and quantification of foot-and-mouth disease virus. Journal of Virological Methods 2020: 113988.

Chen W, Zhang N, Wei J, Yen HL, Li Y. Short-range airborne route dominates exposure of respiratory infection during close contact. Building and Environment 2020a; 176: 106859–106859.

Chen Y, Chen L, Deng Q, Zhang G, Wu K, Ni L, et al. The presence of SARS-CoV-2 RNA in the feces of COVID-19 patients. Journal of medical virology 2020b.

Chia PY, Coleman KK, Tan YK, Ong SWX, Gum M, Lau SK, et al. Detection of air and surface contamination by SARS-CoV-2 in hospital rooms of infected patients. Nature Communications 2020; 11.

Chin AW, Poon LL. Stability of SARS-CoV-2 in different environmental conditions–Authors’ reply. The Lancet Microbe 2020; 1: e146.

Dunn RR, Fierer N, Henley JB, Leff JW, Menninger HL. Home life: factors structuring the bacterial diversity found within and between homes. PloS one 2013; 8: e64133.

Elsamadony M, Fujii M, Miura T, Watanabe T. Possible transmission of viruses from contaminated human feces and sewage: Implications for SARS-CoV-2. Science of the Total Environment 2021; 755: 142575–142575.

Fabian P, McDevitt JJ, Lee WM, Houseman EA, Milton DK. An optimized method to detect influenza virus and human rhinovirus from exhaled breath and the airborne environment. Journal of Environmental Monitoring 2009; 11: 314–317.

Feng B, Xu K, Gu S, Zheng S, Zou Q, Xu Y, et al. Multi-route transmission potential of SARS-CoV-2 in healthcare facilities. Journal of Hazardous Materials 2021; 402: 123771–123771.

Givehchi R, Maestre JP, Bi C, Wylie D, Xu Y, Kinney KA, et al. Quantitative filter forensics with residential HVAC filters to assess indoor concentrations. Indoor air 2019; 29: 390–402.

Gonzalez R, Curtis K, Bivins A, Bibby K, Weir MH, Yetka K, et al. COVID-19 surveillance in Southeastern Virginia using wastewater-based epidemiology. Water Research 2020; 186: 116296–116296.

Goyal SM, Anantharaman S, Ramakrishnan M, Sajja S, Kim SW, Stanley NJ, et al. Detection of viruses in used ventilation filters from two large public buildings. American journal of infection control 2011; 39: e30–e38.

Grijalva C, Rolfes M, Zhu Y, McLean H, Hanson K, Belongia E, et al. Transmission of SARS-COV-2 Infections in Households — Tennessee and Wisconsin. MMWR Morb Mortal Wkly Rep. 2020.

Guillier L, Martin-Latil S, Chaix E, Thébault A, Pavio N, Le Poder S, et al. Modeling the inactivation of viruses from the coronaviridae family in response to temperature and relative humidity in suspensions or on surfaces. Applied and environmental microbiology 2020; 86.

Guo Z-D, Wang Z-Y, Zhang S-F, Li X, Li L, Li C, et al. Aerosol and Surface Distribution of Severe Acute Respiratory Syndrome Coronavirus 2 in Hospital Wards, Wuhan, China, 2020. Emerging Infectious Diseases 2020; 26.

Haaland D, Siegel J. Quantitative filter forensics for indoor particle sampling. Indoor Air 2017; 27: 364–376.

Harris PA, Taylor R, Minor BL, Elliott V, Fernandez M, O’Neal L, et al. The REDCap consortium: Building an international community of software platform partners. Journal of Biomedical Informatics 2019; 95: 103208–103208.

Harris PA, Taylor R, Thielke R, Payne J, Gonzalez N, Conde JG. Research electronic data capture (REDCap)-A metadata-driven methodology and workflow process for providing translational research informatics support. Journal of Biomedical Informatics 2009; 42: 377–381.

Hirotsu Y, Maejima M, Nakajima M, Mochizuki H, Omata M. Environmental cleaning is effective for the eradication of severe acute respiratory syndrome coronavirus 2 (SARS-CoV-2) in contaminated hospital rooms: A patient from the Diamond Princess cruise ship. Infection Control and Hospital Epidemiology 2020: 0–2.

Horve PF, Dietz L, Fretz M, Constant DA, Wilkes A, Townes JM, et al. Identification of SARS-CoV-2 RNA in Healthcare Heating, Ventilation, and Air Conditioning Units. medRxiv 2020.

Iker BC, Bright KR, Pepper IL, Gerba CP, Kitajima M. Evaluation of commercial kits for the extraction and purification of viral nucleic acids from environmental and fecal samples. Journal of virological methods 2013; 191: 24–30.

Kampf G, Todt D, Pfaender S, Steinmann E. Persistence of coronaviruses on inanimate surfaces and their inactivation with biocidal agents. Journal of Hospital Infection 2020; 104: 246–251.

Lescure F-X, Bouadma L, Nguyen D, Parisey M, Wicky P-H, Behillil S, et al. Clinical and virological data of the first cases of COVID-19 in Europe: a case series. The Lancet Infectious Diseases 2020.

Lewis NM, Chu VT, Ye D, Conners EE, Gharpure R, Laws RL, et al. Household Transmission of SARS-CoV-2 in the United States. Clinical Infectious Diseases 2020.

Li W, Zhang B, Lu J, Liu S, Chang Z, Cao P, et al. The characteristics of household transmission of COVID-19. Clinical Infectious Diseases 2020.

Liu F, Cai Z-b, Huang J-s, Niu H-y, Yu W-y, Zhang Y, et al. Repeated COVID-19 relapse during post-discharge surveillance with viral shedding lasting for 67 days in a recovered patient infected with SARS-CoV-2. Journal of Microbiology, Immunology and Infection 2020.

Lopez AS, Hill M, Antezano J, Vilven D, Rutner T, Bogdanow L, et al. Transmission dynamics of COVID-19 outbreaks associated with child care facilities—Salt Lake City, Utah, April–July 2020. Morbidity and Mortality Weekly Report 2020; 69: 1319.

Lu X, Wang L, Sakthivel SK, Whitaker B, Murray J, Kamili S, et al. US CDC real-time reverse transcription PCR panel for detection of severe acute respiratory syndrome coronavirus 2. Emerging infectious diseases 2020; 26: 1654.

Ma J, Qi X, Chen H, Li X, Zhang Z, Wang H, et al. COVID-19 patients in earlier stages exhaled millions of SARS-CoV-2 per hour. Clinical Infectious Diseases 2020.

Madewell ZJ, Yang Y, Longini Jr IM, Halloran ME, Dean NE. Household transmission of SARS-CoV-2: a systematic review and meta-analysis of secondary attack rate. medRxiv 2020.

Maestre JP, Jennings W, Wylie D, Horner SD, Siegel J, Kinney KA. Filter forensics: microbiota recovery from residential HVAC filters. Microbiome 2018; 6: 1–14.

Mahdavi A, Siegel JA. Extraction of Dust Collected in HVAC Filters for Quantitative Filter Forensics. Aerosol Science and Technology 2020: 1–15.

Matson MJ, Yinda CK, Seifert SN, Bushmaker T, Fischer RJ, van Doremalen N, et al. Effect of environmental conditions on SARS-CoV-2 stability in human nasal mucus and sputum. Emerging infectious diseases 2020; 26: 2276.

Miller SL, Nazaroff WW, Jimenez JL, Boerstra A, Buonanno G, Dancer SJ, et al. Transmission of SARS-CoV-2 by inhalation of respiratory aerosol in the Skagit Valley Chorale superspreading event. Indoor air 2020.

Morawska L, Cao J. Airborne transmission of SARS-CoV-2: The world should face the reality. Environment International 2020: 105730.

Mouchtouri VA, Koureas M, Kyritsi M, Vontas A, Kourentis L, Sapounas S, et al. aEnvironmental contamination of SARS-CoV-2 on surfaces, air-conditioner and ventilation systems. International journal of hygiene and environmental health 2020; 230: 113599.

Nissen K, Krambrich J, Akaberi D, Hoffman T, Ling J, Lundkvist Å, et al. Long-distance airborne dispersal of SARS-CoV-2 in COVID-19 wards. Scientific Reports 2020; 10: 19589.

Noris F, Siegel JA, Kinney KA. Evaluation of HVAC filters as a sampling mechanism for indoor microbial communities. Atmospheric Environment 2011; 45: 338–346.

Ong SWX, Tan YK, Chia PY, Lee TH, Ng OT, Wong MSY, et al. Air, Surface Environmental, and Personal Protective Equipment Contamination by Severe Acute Respiratory Syndrome Coronavirus 2 (SARS-CoV-2) from a Symptomatic Patient. JAMA - Journal of the American Medical Association 2020; 323: 1610–1612.

Pan Y, Zhang D, Yang P, Poon LL, Wang Q. Viral load of SARS-CoV-2 in clinical samples. The Lancet Infectious Diseases 2020; 20: 411–412.

Prather KA, Marr LC, Schooley RT, McDiarmid MA, Wilson ME, Milton DK. Airborne transmission of SARS-CoV-2. Science (New York, NY) 2020: eabf0521.

Prussin AJ, Vikram A, Bibby KJ, Marr LC. Seasonal dynamics of the airborne bacterial community and selected viruses in a children’s daycare center. PloS one 2016; 11: e0151004.

Qian H, Miao T, Liu L, Zheng X, Luo D, Li Y. Indoor transmission of SARS-CoV-2. Indoor Air 2020; n/a.

Riddell S, Goldie S, Hill A, Eagles D, Drew TW. The effect of temperature on persistence of SARS-CoV-2 on common surfaces. Virology Journal 2020; 17: 145–145.

Sanekata T, Fukuda T, Miura T, Morino H, Lee C, Maeda K, et al. Evaluation of the antiviral activity of chlorine dioxide and sodium hypochlorite against feline calicivirus, human influenza virus, measles virus, canine distemper virus, human herpesvirus, human adenovirus, canine adenovirus and canine parvovirus. Biocontrol science 2010; 15: 45–49.

Santarpia JL, Rivera DN, Herrera VL, Morwitzer MJ, Creager HM, Santarpia GW, et al. Aerosol and surface contamination of SARS-CoV-2 observed in quarantine and isolation care. Scientific Reports 2020; 10: 1–8.

Shirai J, Kanno T, Tsuchiya Y, Mitsubayashi S, Seki R. Effects of chlorine, iodine, and quaternary ammonium compound disinfectants on several exotic disease viruses. Journal of Veterinary Medical Science 2000; 62: 85–92.

Shrestha PM, Humphrey JL, Barton KE, Carlton EJ, Adgate JL, Root ED, et al. Impact of Low-Income Home Energy-Efficiency Retrofits on Building Air Tightness and Healthy Home Indicators. Sustainability 2019; 11: 2667.

Staudt A, Saunders J, Pavlin J, Shelton-Davenport M. Airborne Transmission of SARS-CoV-2: Proceedings of a Workshop—in Brief. 2020.

Tang M, Zhu N, Kinney K, Novoselac A. Transport of indoor aerosols to hidden interior spaces. Aerosol Science and Technology 2020; 54: 94–110.

To KK-W, Tsang OT-Y, Leung W-S, Tam AR, Wu T-C, Lung DC, et al. Temporal profiles of viral load in posterior oropharyngeal saliva samples and serum antibody responses during infection by SARS-CoV-2: an observational cohort study. The Lancet Infectious Diseases 2020.

Tuladhar E, de Koning MC, Fundeanu I, Beumer R, Duizer E. Different virucidal activities of hyperbranched quaternary ammonium coatings on poliovirus and influenza virus. Applied and environmental microbiology 2012a; 78: 2456–2458.

Tuladhar E, Terpstra P, Koopmans M, Duizer E. Virucidal efficacy of hydrogen peroxide vapour disinfection. Journal of Hospital Infection 2012b; 80: 110–115.

Van Doremalen N, Bushmaker T, Morris DH, Holbrook MG, Gamble A, Williamson BN, et al. Aerosol and surface stability of SARS-CoV-2 as compared with SARS-CoV-1. New England Journal of Medicine 2020; 382: 1564–1567.

Wang Y, Tian H, Zhang L, Zhang M, Guo D, Wu W, et al. Reduction of secondary transmission of SARS-CoV-2 in households by face mask use, disinfection and social distancing: a cohort study in Beijing, China. BMJ Global Health 2020a; 5: e002794.

Wang Z, Ma W, Zheng X, Wu G, Zhang R. Household transmission of SARS-CoV-2. Journal of Infection 2020b.

Wölfel R, Corman VM, Guggemos W, Seilmaier M, Zange S, Müller MA, et al. Virological assessment of hospitalized patients with COVID-2019. Nature 2020; 581: 465–469.

Wolff MH, Sattar SA, Adegbunrin O, Tetro J. Environmental survival and microbicide inactivation of coronaviruses. Coronaviruses with special emphasis on first insights concerning SARS. Springer, 2005, pp. 201–212.

Wu J, Huang Y, Tu C, Bi C, Chen Z, Luo L, et al. Household Transmission of SARS-CoV-2, Zhuhai, China, 2020. Clinical Infectious Diseases 2020.

Xu Y, Li X, Zhu B, Liang H, Fang C, Gong Y, et al. Characteristics of pediatric SARS-CoV-2 infection and potential evidence for persistent fecal viral shedding. Nature Medicine 2020; 26: 502–505.

Yamagishi T, Ohnishi M, Matsunaga N, Kakimoto K, Kamiya H, Okamoto K, et al. Environmental sampling for severe acute respiratory syndrome coronavirus 2 during a COVID-19 outbreak on the diamond princess cruise ship. The Journal of infectious diseases 2020; 222: 1098–1102.

Yamamoto N, Shendell DG, Winer AM, Zhang J. Residential air exchange rates in three major US metropolitan areas: results from the Relationship Among Indoor, Outdoor, and Personal Air Study 1999–2001. Indoor Air 2010; 20: 85–90.

Yang W, Elankumaran S, Marr LC. Concentrations and size distributions of airborne influenza A viruses measured indoors at a health centre, a day-care centre and on aeroplanes. Journal of the Royal Society Interface 2011; 8: 1176–1184.

Ye G, Lin H, Chen S, Wang S, Zeng Z, Wang W, et al. Environmental contamination of SARS-CoV-2 in healthcare premises. Journal of Infection 2020: 2–6.

Zuo Z, de Abin M, Chander Y, Kuehn TH, Goyal SM, Pui DYH. Comparison of spike and aerosol challenge tests for the recovery of viable influenza virus from non-woven fabrics. Influenza and other Respiratory Viruses 2013; 7: 637–644.

