## Supplemental Information for "Distribution of SARS-CoV-2 RNA Signal in a Home with COVID-19 Positive Occupants"

**Supplementary information**

1. RT-qPCR LOD
2. All swab samples in recovered copies per swab
3. Cleaning regime
4. BRSV recovery
5. N1-N2 Comparison
6. Active ingredient in cleaners
7. QFF calculations

### *1. LOD*

The limit of detection (LOD) was determined by using six serial dilutions of SARS-CoV-2 standards (from 100,000 to 1 copy per reaction). After narrowing the initial range, 20 replicates of the standards 10, 5, 2.5 and 1 copy per reaction were analyzed. Following the procedure by Klymus et al. (2019) the effective LOD determined by probit modeling when analyzing samples in triplicate was estimated to be 1.5 copies/ul (CI= 1.13-1.87).

### *2. Recovered copy numbers for all swab samples*

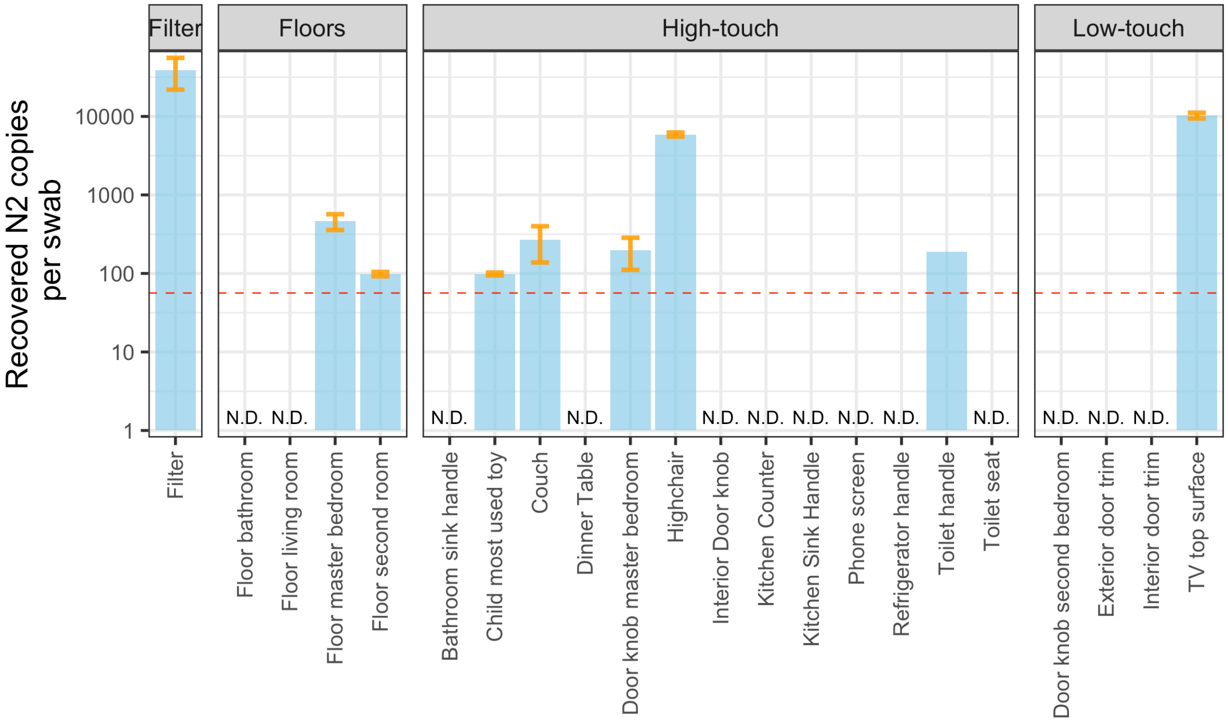


Fig S1. All fomites sampled, results presented in recovered copies per swab, red line represents the effective LOD

***3. Cleaning regime***

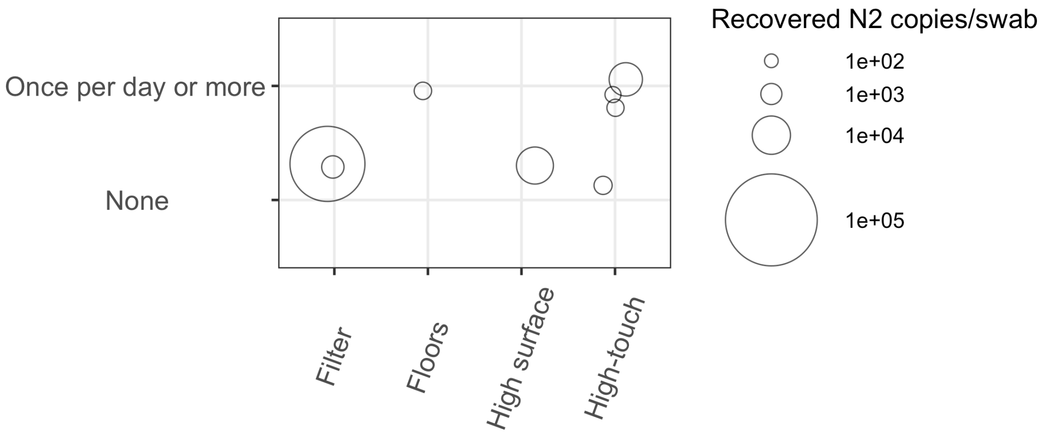


Fig. S2. Recovered copies per swab as a function of the cleaning regime and the sample location.

***4. BRSV recovery***

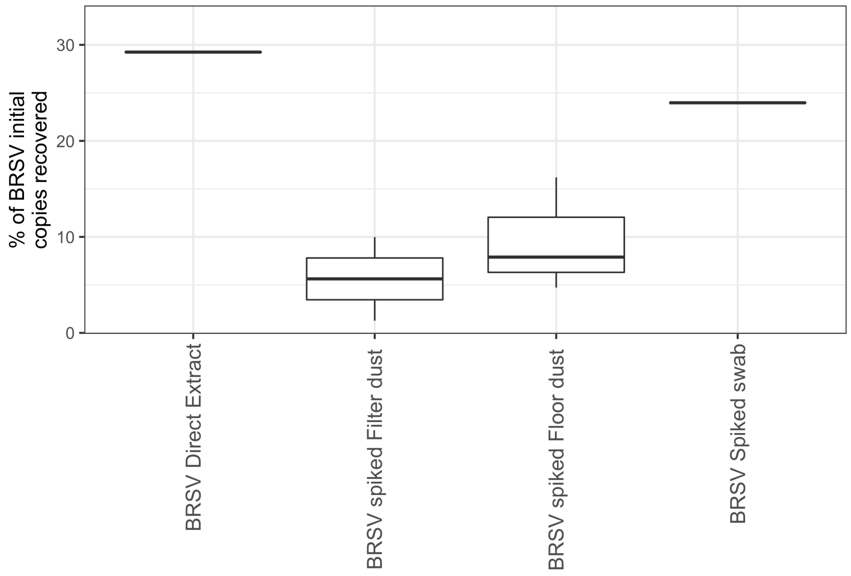


Fig S3. Percentage recovery of BRSV virus.

### *5. N1-N2 Comparison*

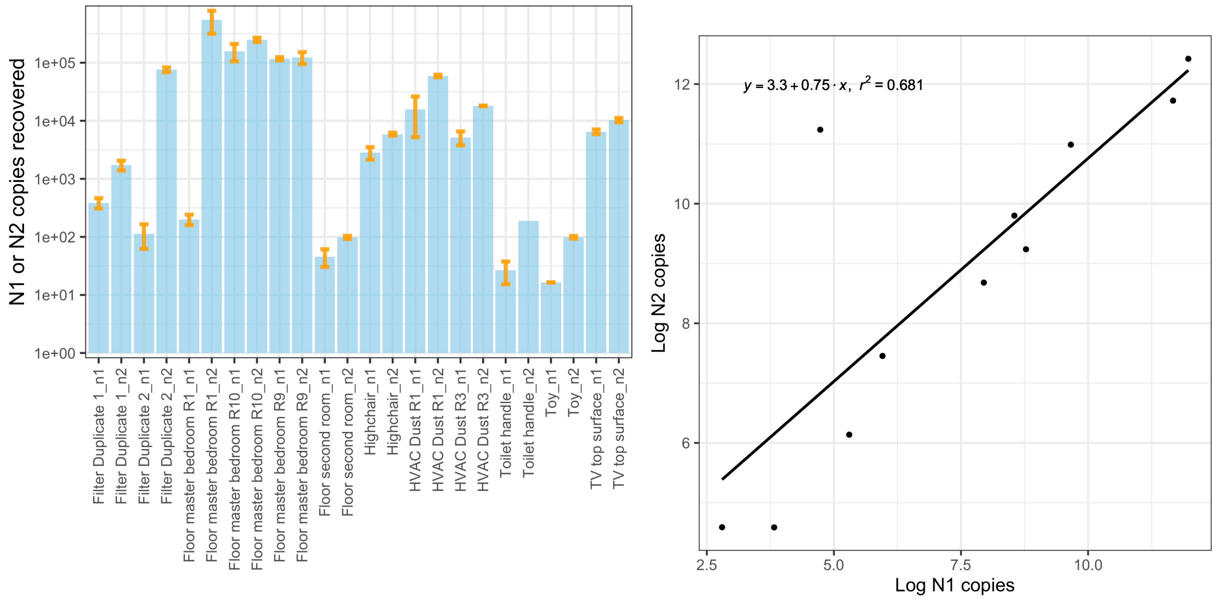


Figure S4. Left) N1-N2 copies recovered per swab across the fomites tested with both primer sets. Right) Correlation between N1-N2

### *6. Active ingredient in cleaners*

Table S1 Cleaner and main active ingredient

| Cleaner | Active ingredients |
|---|---|
| Cleaner 1 | Glycolic Acid |
| Cleaner 2 | Alkyl (C12 [40%]; C14 [50%]; C16 [10%]) Dimethyl Benzyl Ammonium Chloride |
| Cleaner 3 | Sodium Laureth Sulfate |

### 7. *Quantitative Filter Forensics Calculations*

Table S2. Quantitative Filter Forensics calculation of the temporally and spatially integrated airborne concentration of SARS-CoV-2 virus

| Variable | Value | Units | Considerations | Reference |
|---|---|---|---|---|
| m Mass of dust from HVAC filter | 0.68 | g | Includes physical recovery of 27% of the dust by researcher from the filter and 68% efficacy of the participant with respect to researcher | Mahdavi et al., 2020 |
| f Concentration SARS-CoV-2 | 8.58E+06 | N2 copies/g | Includes average 5.5% viral recovery as per BRSV during RNA extraction | This work |
| t Runtime | 240 | h | Median Runtime during summer in a similar area of this study | Givehchi et al., 2019 |
| Q Flowrate | 1350 | $m^3$/h | Median Flowrate during summer in a similar area of this study | Givehchi et al., 2019 |
| η Filtration efficiency estimate | 20 | % | MERV 4 Filter | Siegel., 2020, Personal communication, Persily et al., 2006 |

### References


Givehchi R, Maestre JP, Bi C, Wylie D, Xu Y, Kinney KA, et al. Quantitative filter forensics with residential HVAC filters to assess indoor concentrations. Indoor air 2019; 29: 390-402.

Klymus KE, Merkes CM, Allison MJ, Goldberg CS, Helbing CC, Hunter ME, et al. Reporting the limits of detection and quantification for environmental DNA assays. Environmental DNA 2019.

Mahdavi A, Siegel JA. Extraction of Dust Collected in HVAC Filters for Quantitative Filter Forensics. Aerosol Science and Technology 2020: 1-15.

Persily A, Chapman RE, Emmerich SJ, Dols WS, Davis H, Lavappa P, et al. Building retrofits for increased protection against airborne chemical and biological releases. National Institute of Standards and Technology, Gaithersburg, MD 2007.
